# White Matter β-Amyloid Precursor Protein Immunoreactivity in Autopsied Subjects With and Without COVID-19

**DOI:** 10.1101/2021.12.16.21266656

**Authors:** Thomas G. Beach, Michael DeTure, Jessica E. Walker, Richard Arce, Michael J. Glass, Lucia I. Sue, Anthony J. Intorcia, Courtney M. Nelson, Katsuko E. Suszczewicz, Claryssa I. Borja, Geidy E. Serrano, Dennis W. Dickson

## Abstract

The coronavirus SARS-CoV-2 causes COVID-19, a predominantly respiratory disease that has been reported to be associated with numerous neurological signs, symptoms and syndromes. More than 20 published studies have used RT-PCR methods to determine viral SARS-CoV-2 genomic presence in postmortem brain tissue and the overall impression is that viral brain invasion is relatively uncommon and occurs in low copy numbers, supporting indirect mechanisms as the cause of most neurological phenomena. Hypoxic-ischemic brain injury and stroke are one such possible indirect mechanism, as acute ischemia or stroke concurrence with COVID-19 has been reported as being 0.5% to 20%. Immunohistochemical stains for β-amyloid precursor protein (APP) have been suggested to be a “signature” change of hypoxic leukoencephalopathy or COVID-19 brain disease, although prior reports have not had a non-COVID-19 control group. We therefore compared the prevalence and intensity of white matter APP staining in the brains of subjects dying with and without COVID-19. Clinical and neuropathological results, including semi-quantitative assessment of the density of white matter APP staining, were compared between 20 COVID-19 cases and 20 pre-COVID-19 autopsy cases, including 10 cases with autopsy-proven non-COVID-19 pneumonia and 10 cases without pneumonia. Positive APP white matter staining in at least one of the two brain regions (precentral gyrus and cingulate gyrus) studied was not significantly more common in COVID-19 vs controls (14/20 vs 12/20). Comparing density scores from both brain regions combined, the mean scores for COVID-19 cases were higher than those for controls of both types together but not significantly different when restricting to controls with pneumonia. Among control cases, cases with pneumonia had significantly higher scores. The presence or absence of a major neuropathologically-defined neurodegenerative disorder did not significantly affect the APP scores. The major finding is that while APP white matter staining cannot be regarded as a specific marker of COVID-19, as it does not occur with significantly greater probability in in COVID-19 brains as compared to non-COVID-19 brains, it is possible that white matter APP staining, representing acute or subacute axonal damage, may be a common occurrence in the perimortem period, and that it may be more intense in subjects dying with pneumonia, regardless of cause.

## INTRODUCTION

The coronavirus SARS-CoV-2 (SCV2) has been reported to be associated with numerous neurological signs, symptoms and syndromes, affecting up to 36% of patients ^1-10^. It is not yet determined whether these are due to direct viral action, indirect immune-mediated mechanisms, or to other systemic reactions common to many types of critical illness. COVID-19 is known to be accompanied by coagulopathy, sepsis, autoimmune attack and multiorgan failure^11^. More than 20 published studies have used RT-PCR methods to determine SCV2 genomic presence in postmortem brain tissue ^11-31^ and the overall impression is that SCV2 brain invasion is relatively uncommon and that viral copy numbers are generally low, supporting indirect mechanisms as the cause of most neurological phenomena.

Stroke is a disastrous outcome of COVID-19 disease but how often it occurs is difficult to ascertain due to a wide range in rate estimates. Acute ischemia or stroke concurrence with COVID-19 has been reported as between 0.5% to 20% while rates of acute brain hemorrhage range from 0.13% to 9.5% ^4,5,11,17,18,20,23,25-27,29,30,32-54^. To date, however, most reports have not had a non-COVID-19 control group. A few clinical studies have compared stroke rates in hospitalized COVID-19 and influenza subjects, finding low but slightly increased rates with COVID-19 ^49,50^ (1.6% vs 0.2% and 0.9% vs 0.3%) or alternatively, equivalent rates ^32^ (1.2% for both). We found, in a study of 691 pre-COVID-19 autopsies, that the prevalence of acute ischemia or infarction did not differ between autopsy-confirmed pneumonia and non-pneumonia cases, both being about 14% ^55^. We suggest that COVID-19 and non-COVID-19 pneumonia may have similar risks for concurrent acute brain infarction when pneumonia is severe enough to cause death. Moreover, acute brain ischemia, infarction or hemorrhage may not be more common in subjects dying of acute pneumonia than in subjects dying without acute pneumonia.

These studies, however, have been based on clinical reports or standard autopsy diagnostic methods, and it is possible that more sensitive methods might be more informative. In particular, immunohistochemical stains for β-amyloid precursor protein (APP) have been suggested to be a “signature” change of hypoxic leukoencephalopathy or COVID-19 brain disease ^39,42^. We therefore compared the prevalence and intensity of white matter APP staining in the brains of subjects dying with and without COVID-19.

## MATERIALS AND METHODS

### Human Subjects and Characterization

The COVID-19 subjects were derived from Banner Sun Health Research Institute (BSHRI) in Sun City, Arizona (n =10), or from the Mayo Clinic in Jacksonville, Florida (n = 10). All subjects had been consented for autopsy and subsequent research studies with approval by Institutional Review Boards (Western IRB # 1132516; Mayo Clinic Florida Brain Bank IRB # 15-009452).

Clinical and neuropathological results for the 20 COVID-19 cases are described in our prior publication ^56^ and are briefly re-summarized in Table 1. All COVID-19 subjects had positive clinical diagnostic test results for SCV2 and all were considered to have died in 2020 as a result of severe COVID-19. An additional 20 cases were chosen from 2018 and 2019 pre-COVID-19 BSHRI autopsies, including 10 cases with autopsy-proven non-COVID-19 pneumonia (all with acute bronchopneumonia) and 10 cases without pneumonia.

**Table 1.**
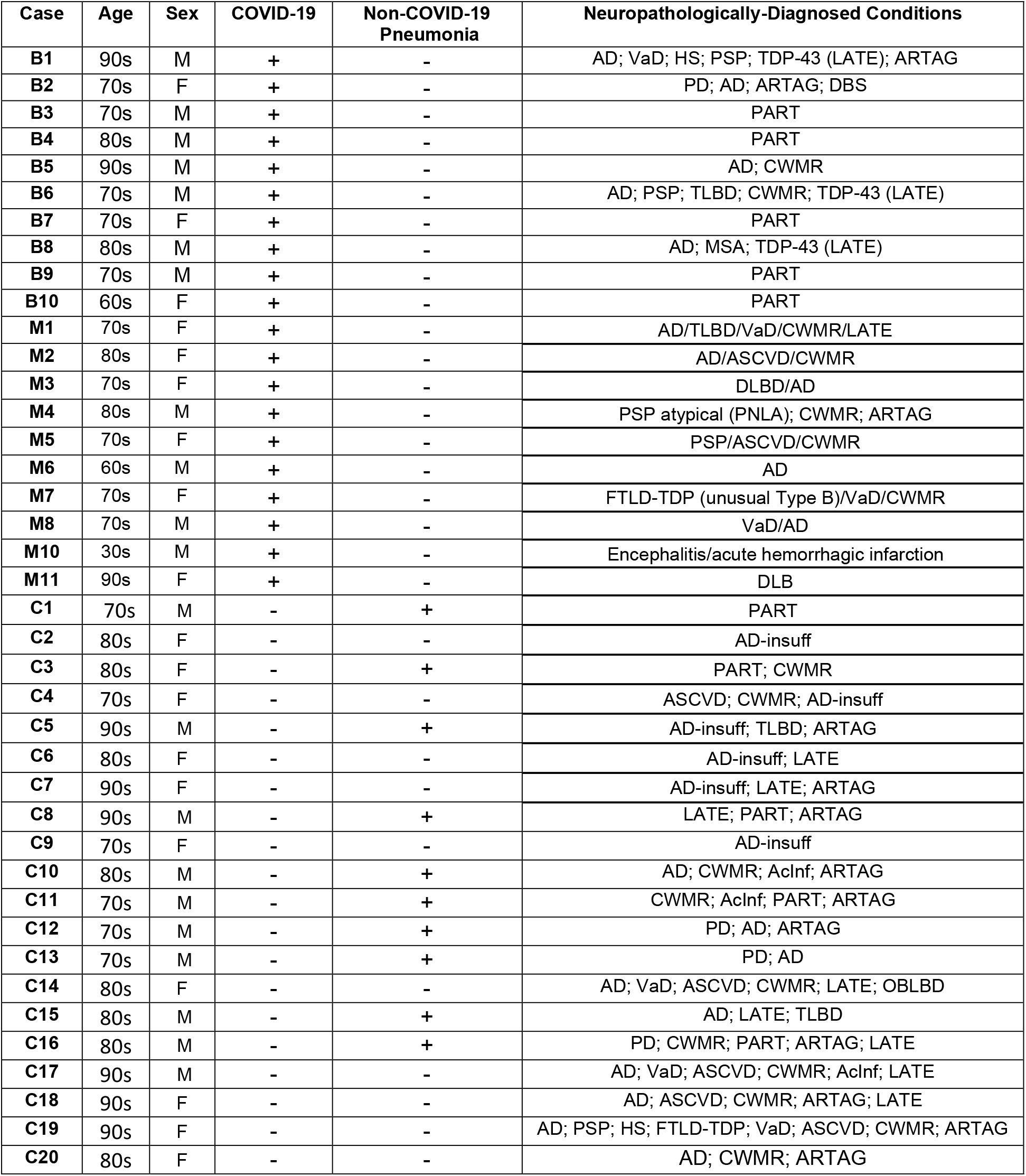
Case data for neuropathologically-diagnosed comorbidities. B = COVID-19 cases derived from BSHRI; M = COVID-19 cases derived from Mayo Clinic; C = Non-COVID-19 control cases derived from BSHRI; AcInf = Acute infarctions; AD = Alzheimer’s disease; AD-insuff = AD changes insufficient for diagnosis; ARTAG = age-related tau astrogliopathy; ASCVD = atherosclerotic cerebrovascular disease; BSLBD = brainstem Lewy body disease; CWMR = cerebral white matter rarefaction; DBS = deep brain stimulator; DLBD = diffuse Lewy body disease; FTLD-TDP = frontotemporal lobar degeneration with TDP-43 proteinopathy; HS = hippocampal sclerosis; LATE = Limbic age-related TDP-43 proteinopathy; OBLBD = olfactory bulb Lewy body disease; PD = Parkinson’s disease; PART = primary age-related tauopathy; PNLA = pallido-nigro-Luysian-degeneration; PSP = progressive supranuclear palsy; TLBD = transitional Lewy body disease; VaD = vascular dementia. Specific ages not given to preserve privacy.

| Case | Age | Sex | COVID-19 | Non-COVID-19<br>Pneumonia | Neuropathologically-Diagnosed Conditions |
| --- | --- | --- | --- | --- | --- |
| B1 | 90s | M | + | - | AD; VaD; HS; PSP; TDP-43 (LATE); ARTAG |
| B2 | 70s | F | + | - | PD; AD; ARTAG; DBS |
| B3 | 70s | M | + | - | PART |
| B4 | 80s | M | + | - | PART |
| B5 | 90s | M | + | - | AD; CWMR |
| B6 | 70s | M | + | - | AD; PSP; TLBD; CWMR; TDP-43 (LATE) |
| B7 | 70s | F | + | - | PART |
| B8 | 80s | M | + | - | AD; MSA; TDP-43 (LATE) |
| B9 | 70s | M | + | - | PART |
| B10 | 60s | F | + | - | PART |
| M1 | 70s | F | + | - | AD/TLBD/VaD/CWMR/LATE |
| M2 | 80s | F | + | - | AD/ASCVD/CWMR |
| M3 | 70s | F | + | - | DLBD/AD |
| M4 | 80s | M | + | - | PSP atypical (PNLA); CWMR; ARTAG |
| M5 | 70s | F | + | - | PSP/ASCVD/CWMR |
| M6 | 60s | M | + | - | AD |
| M7 | 70s | F | + | - | FTLD-TDP (unusual Type B)/VaD/CWMR |
| M8 | 70s | M | + | - | VaD/AD |
| M10 | 30s | M | + | - | Encephalitis/acute hemorrhagic infarction |
| M11 | 90s | F | + | - | DLB |
| C1 | 70s | M | - | + | PART |
| C2 | 80s | F | - | - | AD-insuff |
| C3 | 80s | F | - | + | PART; CWMR |
| C4 | 70s | F | - | - | ASCVD; CWMR; AD-insuff |
| C5 | 90s | M | - | + | AD-insuff; TLBD; ARTAG |
| C6 | 80s | F | - | - | AD-insuff; LATE |
| C7 | 90s | F | - | - | AD-insuff; LATE; ARTAG |
| C8 | 90s | M | - | + | LATE; PART; ARTAG |
| C9 | 70s | F | - | - | AD-insuff |
| C10 | 80s | M | - | + | AD; CWMR; AcInf; ARTAG |
| C11 | 70s | M | - | + | CWMR; AcInf; PART; ARTAG |
| C12 | 70s | M | - | + | PD; AD; ARTAG |
| C13 | 70s | M | - | + | PD; AD |
| C14 | 80s | F | - | - | AD; VaD; ASCVD; CWMR; LATE; OBLBD |
| C15 | 80s | M | - | + | AD; LATE; TLBD |
| C16 | 80s | M | - | + | PD; CWMR; PART; ARTAG; LATE |
| C17 | 90s | M | - | - | AD; VaD; ASCVD; CWMR; AcInf; LATE |
| C18 | 90s | F | - | - | AD; ASCVD; CWMR; ARTAG; LATE |
| C19 | 90s | F | - | - | AD; PSP; HS; FTLTDP; VaD; ASCVD; CWMR; ARTAG |
| C20 | 80s | F | - | - | AD; CWMR; ARTAG |

For both BSHRI and Mayo Clinic, published diagnostic clinicopathological consensus criteria ^57-69^ were used when applicable, incorporating research clinical assessment results as well as pertinent private medical history. The histological sampling and staining incorporated the protocols recommended by the National Institute on Aging and Alzheimer’s Association (NIA-AA) ^67-69^.

Immunohistochemical staining for APP was performed as previously published ^70^ on sections of precentral and cingulate gyrus with underlying white matter. The extent of APP staining was semi-quantitatively graded on a scale of 0-3. To be considered positive there had to be not only increased APP immunoreactivity within axons, but also evidence of focal axonal swelling with or without fragmentation.

Statistical methods included unpaired, two-way t-tests for continuous variables, Kruskal-Wallis and Mann-Whitney U-tests for ordinal variables and Fisher Exact tests for proportions. The significance level was set at p< 0.05.

## RESULTS

Table 1 shows basic data for the cases studied. COVID-19 cases were younger (77.5; SD 2.9) than non-COVID-19 cases (84.0; SD 2.0) but the difference was not significant (p = 0.07). Males made up 11/20 and 10/20 of the COVID-19 and non-COVID-19 cases, respectively (ns). Both COVID-19 and non-COVID-19 cases had a wide range of neuropathological diagnoses, consistent with their age and derivation from academic centers devoted to the study of neurodegenerative disease.

Table 2 and Figure 1 show results of the APP staining in COVID-19 and non-COVID-19 control cases. Positive APP white matter staining was seen in at least one of the two brain regions in 14/20 COVID-19 cases and in 12/20 of the non-COVID-19 control cases (ns). Cases were positive in the precentral gyrus in 11/20 COVID-19 and 5/20 control cases (ns) while in the cingulate gyrus the ratios were 12/20 and 10/20, respectively (ns).

**Table 2.** Results of semi-quantitation (scale of 0–3) of APP staining in COVID-19 and non-COVID-19 control cases. PRCG = precentral gyrus; CING = cingulate gyrus; CWMR = cerebral white matter rarefaction.

| Case | COVID-19 | Non-COVID-19<br>Pneumonia | APP PRCG | APP CING |
| --- | --- | --- | --- | --- |
| B1 | + | - | 0 | 0 |
| B2 | + | - | 1 | 1 |
| B3 | + | - | 0 | 2 |
| B4 | + | - | 1 | 1 |
| B5 | + | - | 3 | 3 |
| B6 | + | - | 0 | 0 |
| B7 | + | - | 0.5 | 0 |
| B8 | + | - | 3 | 0 |
| B9 | + | - | 1 | 1 |
| B10 | + | - | 0 | 0 |
| M1 | + | - | 0.5 | 1 |
| M2 | + | - | 1 | 1.5 |
| M3 | + | - | 0 | 2 |
| M4 | + | - | 0 | 0 |
| M5 | + | - | 0 | 0 |
| M6 | + | - | 2 | 2 |
| M7 | + | - | 0 | 0 |
| M8 | + | - | 0 | 1 |
| M10 | + | - | 2 | 3 |
| M11 | + | - | 1 | 2 |
| C1 | - | + | 1 | 1 |
| C2 | - | - | 0 | 2 |
| C3 | - | + | 0.5 | 2 |
| C4 | - | - | 0 | 0 |
| C5 | - | + | 0 | 2 |
| C6 | - | - | 0 | 0.5 |
| C7 | - | - | 0 | 0 |
| C8 | - | + | 0 | 0 |
| C9 | - | - | 0 | 0 |
| C10 | - | + | 2 | 0 |
| C11 | - | + | 0 | 0.5 |
| C12 | - | + | 0 | 0.5 |
| C13 | - | + | 0 | 1 |
| C14 | - | - | 0 | 0 |
| C15 | - | + | 0 | 0.5 |
| C16 | - | + | 1 | 0 |
| C17 | - | - | 1 | 2 |
| C18 | - | - | 0 | 0 |
| C19 | - | - | 0 | 0 |
| C20 | - | - | 0 | 0 |

**Figure 1.**
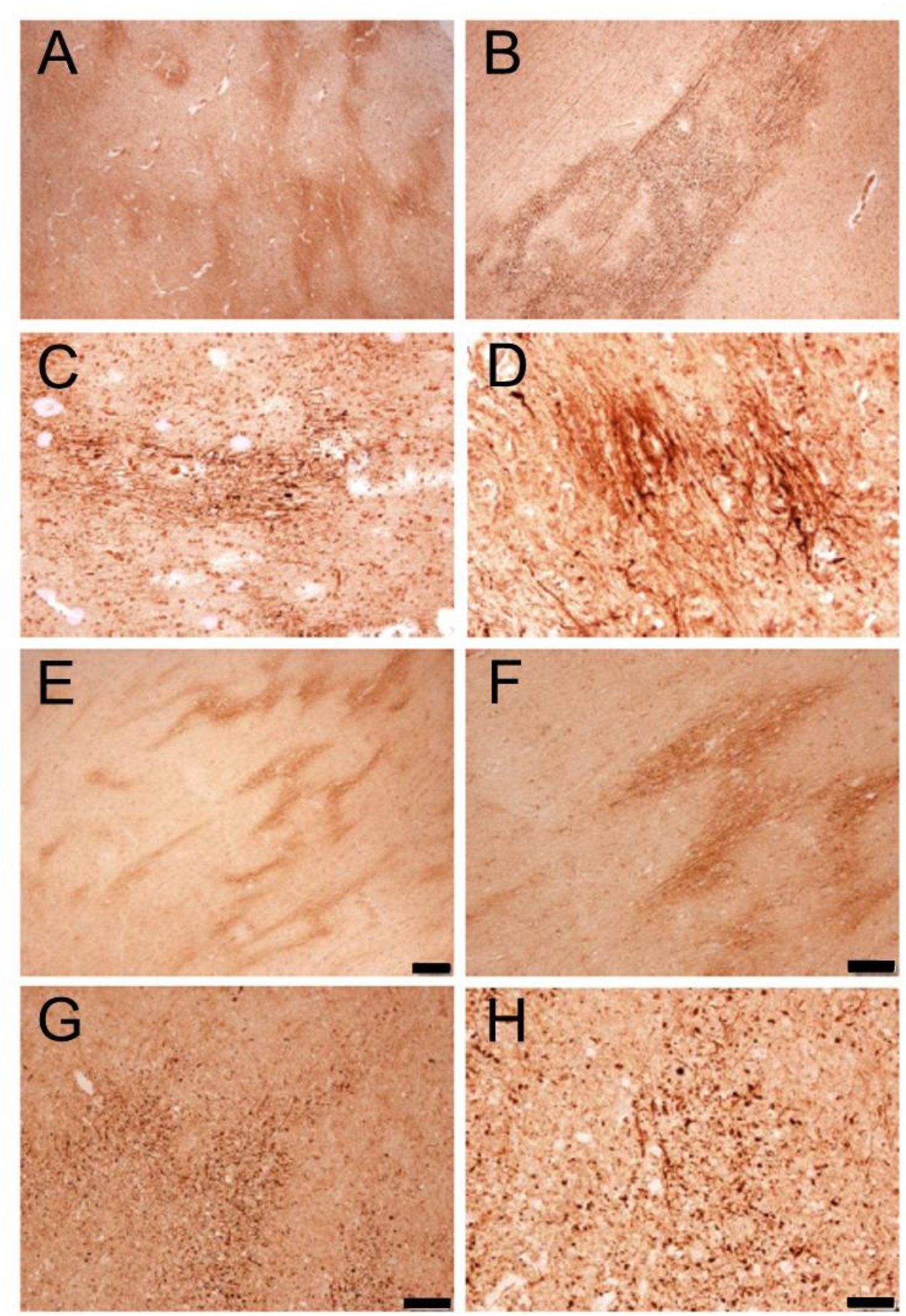
Photomicrographs of cingulate gyrus and precentral gyrus white matter APR staining. A. Cingulate gyrus white matter in Case B9, a man in his 60s with COVID-19. B. Precentral gyrus white matter in Case B8, a man in his 80s with COVID-19. C. Precentral gyrus white matter in Case B5, a man in his 90s with COVID-19. D. Cingulate gyrus white matter in Case B3, a woman in her 70s with COVID-19. E. Cingulate gyrus white matter in Case C5, a man in his 90s with non-COVID-19 pneumonia. F. Precentral gyrus white matter in Case C5, a man in his 90s with non-COVID-19 pneumonia. G. Precentral gyrus white matter in Case C10, a man in his 80s with non-COVID-19 pneumonia. H. Cingulate gyrus white matter in Case C10, a man in his 80s with non-COVID-19 pneumonia. Bar in E also serves for A = 200 um. Bar in F also serves for B = 100 um. Bar in G also serves for C = 50 um. Bar in H also serves for D = 50 um.

Comparing density scores from both brain regions combined, the mean scores for COVID-19 cases were higher than those for controls of both types together (0.91 vs 0.44, Mann-Whitney p = 0.026) but not significantly different (0.91 vs 0.60, p = 0.35) when restricting to controls with pneumonia. Region-specific pairwise scores were not significantly different between COVID-19 and controls of both types (Kruskal-Wallis analysis of variance with subsequent Dunn’s Multiple Comparisons pair-wise testing). In both brain regions, scores were not significantly different between BSHRI and Mayo Clinic COVID-19 cases. Among control cases, when considering both brain regions together, cases with pneumonia had significantly higher scores (0.60 vs 0.27, Mann-Whitney p = 0.044).

When cases were divided by presence (those with AD, VaD, HS, PSP, PD, MSA, DLB, DLBD, or FTLD-TDP in Table 2) or absence of a major neuropathologically-defined neurodegenerative disorder, there were no significant differences in APP scores between the two groups, either when restricting to one brain region or when scores from both brain regions were combined.

When APP scores from both areas were combined, males had significantly greater scores than females (0.89 vs 0.43, Mann-Whitney p = 0.032). There was no significant correlation (Spearman) between age and APP scores for either brain region.

## DISCUSSION

Immunohistochemical staining of swollen white matter axons for β-amyloid precursor protein (APP) has been suggested to be a “signature” change of hypoxic leukoencephalopathy or COVID-19 brain disease ^39,42^. However, these APP-positive features have also been reported in association with a variety of conditions, including human cases and/or animal models of ischemia ^71,72^, traumatic brain injury ^73-75^, Binswanger’s disease or vascular dementia ^76,77^, bacterial meningitis ^78^, drug abuse ^79^ and acute demyelinating diseases ^80^. This investigation examined APP staining in the white matter of 20 subjects that died with COVID-19, in comparison with 20 subjects that died prior to the COVID-19 pandemic. The major finding is that APP white matter staining cannot be regarded as a specific marker of COVID-19 as it does not occur with significantly greater probability in in COVID-19 brains as compared to non-COVID-19 brains. Semi-quantitation of the APP staining gives significantly greater scores for COVID-19 brains as compared to non-COVID-19 brains, but not significantly greater relative to the non-COVID-19 subset that died with autopsy-verified pneumonia.

We did not find greater amounts of APP staining in subjects with major neurodegenerative diseases. Males had greater APP staining scores than females. There was no correlation of APP scores with age.

Based on these findings, it is possible that white matter APP staining, representing acute or subacute axonal damage, may be a common occurrence in the perimortem period, and that it may be more intense in subjects dying with pneumonia, regardless of cause.

Biospecimens from the Banner Sun Health Research Institute Brain and Body Donation Program, including those presented in this report, are available to qualified researchers upon request from https://www.brainandbodydonationregistration.org/.

## Data Availability

All data produced in the present work are contained in the manuscript.

## ACKNOWLEDGEMENTS

This project was supported by a Covid-19 Supplement to a National Institute on Aging grant, (3P30AG019610-20S1), submitted in response to a Notice of Special Interest (NOSI) issued by the National Institute on Aging (NOT-AG-20-022), “to highlight the urgent need for research on Coronavirus Disease 2019…”.

